# Administration of *Bifidobacterium animalis* subsp. *lactis* Strain BB-12^®^ in Healthy Children: Characterization, Functional Composition, and Metabolism of the Gut Microbiome

**DOI:** 10.1101/2023.02.02.23285145

**Authors:** Carlotta Vizioli, Rosario Jaime-Lara, Scott G. Daniel, Alexis Franks, Ana F. Diallo, Kyle Bittinger, Tina P. Tan, Daniel J. Merenstein, Brianna Brooks, Paule V. Joseph, Katherine A. Maki

**Affiliations:** National Institute of Neurological Disease and Stroke, National Institutes of Health, Department of Health and Human Services, Bethesda, MD; National Institute on Alcohol Abuse and Alcoholism National Institutes of Health, Department of Health and Human Services, Bethesda, MD; National Institute of Nursing Research, National Institutes of Health, Department of Health and Human Services, Bethesda, MD; UCLA School of Nursing, University of California Los Angeles, Los Angeles, CA; Division of Gastroenterology, Hepatology, and Nutrition, Children’s Hospital of Philadelphia, Philadelphia, PA; Institute of Inclusion, Inquiry & Innovation (iCubed), Family and Community Health Nursing, School of Nursing, Virginia Commonwealth University, Richmond, VA; Department of Family Medicine, Georgetown University Medical Center, Washington, DC; Translational Biobehavioral and Health Disparities Branch, National Institutes of Health, Clinical Center, Bethesda, MD, 20814

**Keywords:** probiotics, gut microbiome, *L. delbrueckii*, B. animalis BB-12, S. *thermophilus*, metagenomics, metabolomics, children

## Abstract

The consumption of probiotics may influence children’s gut microbiome and metabolome, which may reflect shifts in gut microbial diversity composition and metabolism. These potential changes might have a beneficial impact on health. However, there is a lack of evidence investigating the effect of probiotics on the gut microbiome and metabolome of children. We aimed to examine the potential impact of a two (*Streptococcus thermophilus* and *Lactobacillus delbrueckii*; S2) *vs*. three (S2 + *Bifidobacterium animalis* subsp. *lactis* strain BB-12) strain-supplemented yogurt. Included in this study were 59 participants, aged one to five years old, recruited to phase I of a double-blinded, randomized controlled trial. Fecal samples were collected at baseline, after the intervention, and at twenty days post-intervention discontinuation, and untargeted metabolomics and shotgun metagenomics were performed. Shotgun metagenomics and metabolomic analyses showed no global changes in either intervention group’s gut microbiome alpha or beta diversity indices. The relative abundance of the two and three intervention bacteria increased in the S2 and S2 + BB12 groups, respectively, from *Day 0* to *Day 10*. In the S2+BB12 group, the abundance of several fecal metabolites was reduced at *Day 10*, including alanine, glycine, lysine, phenylalanine, serine, and valine. These fecal metabolite changes did not occur in the S2 group. Future research using longer probiotic intervention durations and in children at risk for gastrointestinal disorders may elucidate if functional metabolite changes confer a protective gastrointestinal effect.

## Introduction

The gut microbiome is comprised of the entire gastrointestinal (GI) microbial community, including bacteria, fungi, viruses, and their genes. Metagenomic analysis captures a comprehensive summary of the microbiome, *i*.*e*., microbial diversity and their ecological niches (microbial function). [1] Gut colonization starts prenatally and continues after birth. The gut microbiome in early infancy begins to stabilize early in life. [2] Several mechanisms, including birth mode, [3] type of milk received, [4] and environmental factors, [5] shape the development of the gut microbiome from infancy to adulthood. The environment and diet during the first two to five years turn an immature microbiota into a more stable, resilient, adult-like gut microbial community. [5] The human gut microbiome influences nutritional absorption, immune health, and behavior. [6] Pre-clinical and clinical studies suggest that the gut microbiota-immune system crosstalk may be responsible for long-term health. [7, 8] Disruptions to a healthy gut microbiome are observed during disease states and across chronic illnesses, including inflammatory and immune disorders. [9-11]

Probiotics are defined by the International Scientific Association for Probiotics and Prebiotics as “live microorganisms that, when administered in adequate amounts, confer a health benefit on the host”. [12] Probiotics are increasingly used in commercial products because of their potential benefits on the gut microbiota that have been shown to exert positive effects on host physiology. [12, 13] The mechanism of action by which probiotics confer health benefits are diverse and include: colonization and normalization of perturbed intestinal microbial populations, competitive exclusion of pathogens, and modulation of the immune system via production of anti-inflammatory factors. [14]

Probiotics have been used in treatment of GI symptoms and prevention or management of GI disorders. Several studies have shown that probiotic strains from the *Bifidobacterium* genus promote the growth of beneficial bacteria, inhibit pathogenic microorganisms by secreting antibacterial factors, [15] competitive adhesion to intestinal epithelial cells [14] improve GI barrier, [16, 17] promoting the formation of mucous layers maintaining of intestinal immune homeostasis, [18] and lower inflammatory cytokines. [19] Moreover, consumption of *Lactobacillus* and *Bifidobacterium* genera have been associated with improved mental health and memory function in pre-clinical and human studies. [20-23]

There is increasing interest in using probiotics as a tool to maintain and restore a healthy gut microbiota. While evidence supports their use in some GI diseases, the impact of probiotics on healthy gut microbiota and its metabolism is still unclear in both adults and children. [24-27] Few studies have examined the effect of probiotics in healthy adults, [26-28] and even fewer have studied the effect of probiotics in healthy children. [29] Although *Bifidobacterium animalis* subsp. *lactis* BB-12 (BB-12) is among the most common probiotic supplements and has previously been demonstrated to be well-tolerated by healthy children, [30] there are limited studies examining the effects of BB-12 on structural and functional characteristics of the gut microbiome in children ages one to five years old. [30]

Metabolites produced by gut microorganisms have been identified in modulating human health, including the immune system, metabolic, and neurobehavioral traits. [14, 21, 28, 31, 32] Furthermore, emerging studies suggest that probiotics’ effects on intestinal metabolites may contribute to intestinal health and immune function. [32-36] Thus, there is growing interest in the link between probiotic administration and the subsequent impact on metabolite changes in the context of human health and disease. Shotgun metagenomics sequencing and untargeted metabolomics technologies have grown exponentially in the last decade providing a key tool to closer examine the microbial characterization, function, and metabolism in a sample (*e*.*g*., fecal or tissue samples). Therefore, metabolomics offers an efficient and accurate strategy of exploring the biological role of how probiotics may impact the pediatric gut microbiome, including how these metabolites respond to different combinations of symbiotic bacteria administration (i.e., *Bifidobacterium vs. Lactobacillus* spp. probiotic genera). [37]

Exploring the effect of probiotics on the gut microbiome and metabolome in healthy children may provide more extensive insight into the relationship between probiotics, gut microbiota, metabolites, and human health. This may aid in developing more effective methods of assessing gut health by simultaneously characterizing the gut microbiome and functional impacts of microbiome community changes on the metabolome. Furthermore, GI disorders are among the most common ailments reported in pediatric primary care. Therefore, utilizing our approach including microbiome and metabolome analyses to characterize structural and functional responses of gut microbiota to two probiotics combinations (*Streptococcus thermophilus* and *Lactobacillus delbrueckii* [S2] *vs. Streptococcus thermophilus, Lactobacillus delbrueckii*, and *Bifidobacterium animalis* subsp. *lactis* strain BB-12 [S2+BB12]) in children is an important first step to design future research evaluating the efficacy of probiotics in the prevention of dysbiosis–associated GI disorders.

To date, few studies have been conducted to examine the effect of probiotics on gut microbial compositional and functional structure combined with associated fecal metabolome changes in healthy children. Thus, this study uses shotgun metagenomic sequencing and untargeted fecal metabolomics to examine the effects induced by the consumption of yogurt with and without the BB-12 probiotic strain (BB12) in healthy children aged one to five years old. This study builds upon findings by Tan et al. [30] using shotgun metagenomics sequencing for microbiome analysis and the integration of metabolomic data.

## Results

### Participant Characteristics

A total of 59 healthy participants between the ages of one and five years (mean age =2.38±1.22) were included in the study (Figure 1A). The yogurt was administered to the participants for ten consecutive days. Fecal samples were analyzed at baseline (*Day 0*), ten (*Day 10*), and 30 (*Day 30*) days from both S2+BB12 (n = 25, 28, and 25, respectively) and S2 (n = 31, 30, and 30, respectively) groups (Figure 1B).

**Figure 1:**
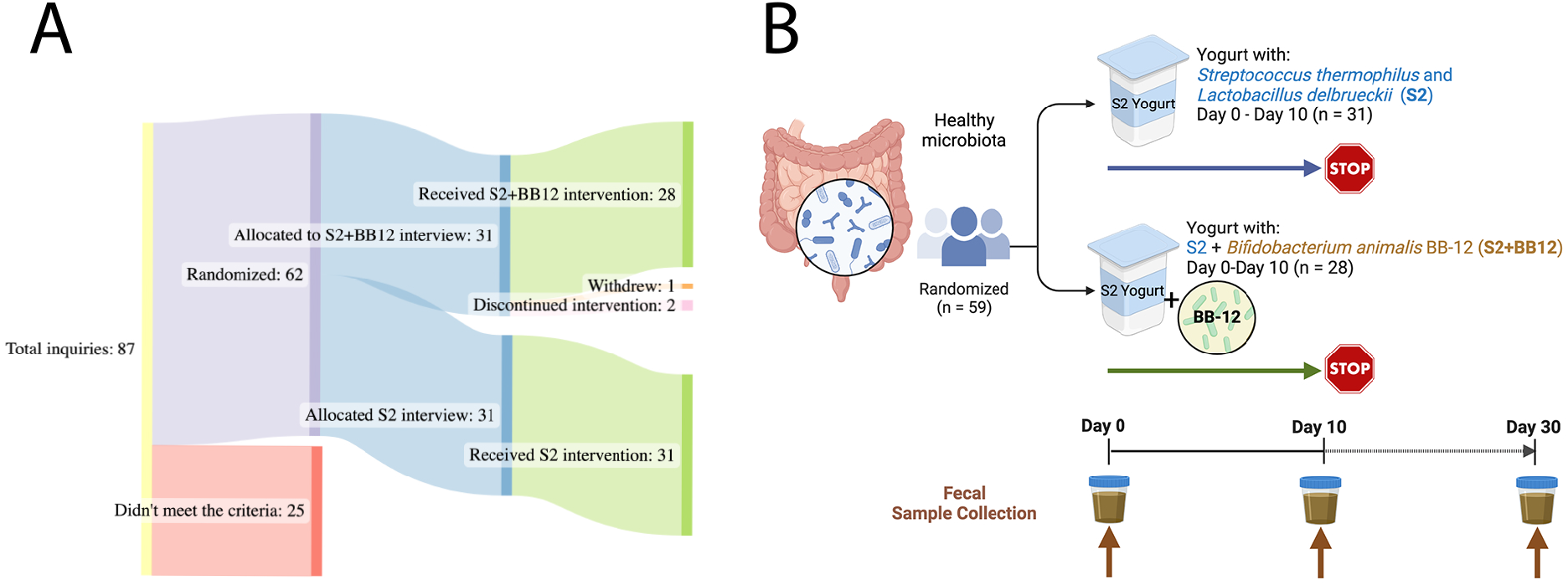
Study Design: A total of 59 children, aged one to five years old, were analyzed for this double-blinded, randomized controlled study (A). The participants consumed yogurt with two (S2 group) *vs*. three (S2+BB12 group) probiotics’ strains (B). The participants consumed yogurt during ten consecutive days. Fecal samples were collected before the intervention (*Day 0*), after ten days of yogurt consumption (*Day 10*), and after ten days of yogurt consumption discontinuation (*Day 30*).

Participants were relatively divided equally between males and females (n = 28 males and n = 31 females). Most of the participants included were White (n = 40). Additional demographic information from the included participants is displayed in Table 1.

**Table 1.** Participants demographics

|  |  | Overall<br>n=59 | S2<br>n=31 | S2+BB12<br>n=28 |
| --- | --- | --- | --- | --- |
| Age | Mean (SD) | 2.38<br>(1.22) | 2.42<br>(1.21) | 2.35<br>(1.26) |
| Gender (n) | Male | 28 | 13 | 15 |
|  | Female | 31 | 18 | 13 |
| Race (n) | Indian/Alaskan | 1 |  | 1 |
|  | Black or African | 8 | 4 | 4 |
|  | White/Caucasian | 40 | 17 | 23 |
|  | Other/more than one race | 7 | 6 | 1 |
| Ethnicity (n) | Hispanic/Latino | 8 | 5 | 3 |

### Metagenomic analyses

The administration of BB-12, in addition to *S. thermophilus* and *L. delbrueckii*, influenced the composition of the gut microbiome, although there were no overt global microbiome changes quantified by alpha and beta diversity indices. Species richness and Shannon indices were not significantly different between the S2 and S2+BB12 groups at *Day 10* (*p* = .565 and *p* = .462, respectively) or *Day 30* (*p* = .669 and *p* = .104, respectively; Figure 2A-B). Similarly, there were no group differences in beta diversity (based on Bray-Curtis dissimilarity) at *Day 10* (R^2^ = 0.02, *p* = .318; Figure 2C) or *Day 30* (R^2^ = 0.02, *p* = .3537; Figure S1). These findings agree with previously reported results by Tan et al. using 16S rRNA analysis **[30]**.

**Figure 2:**
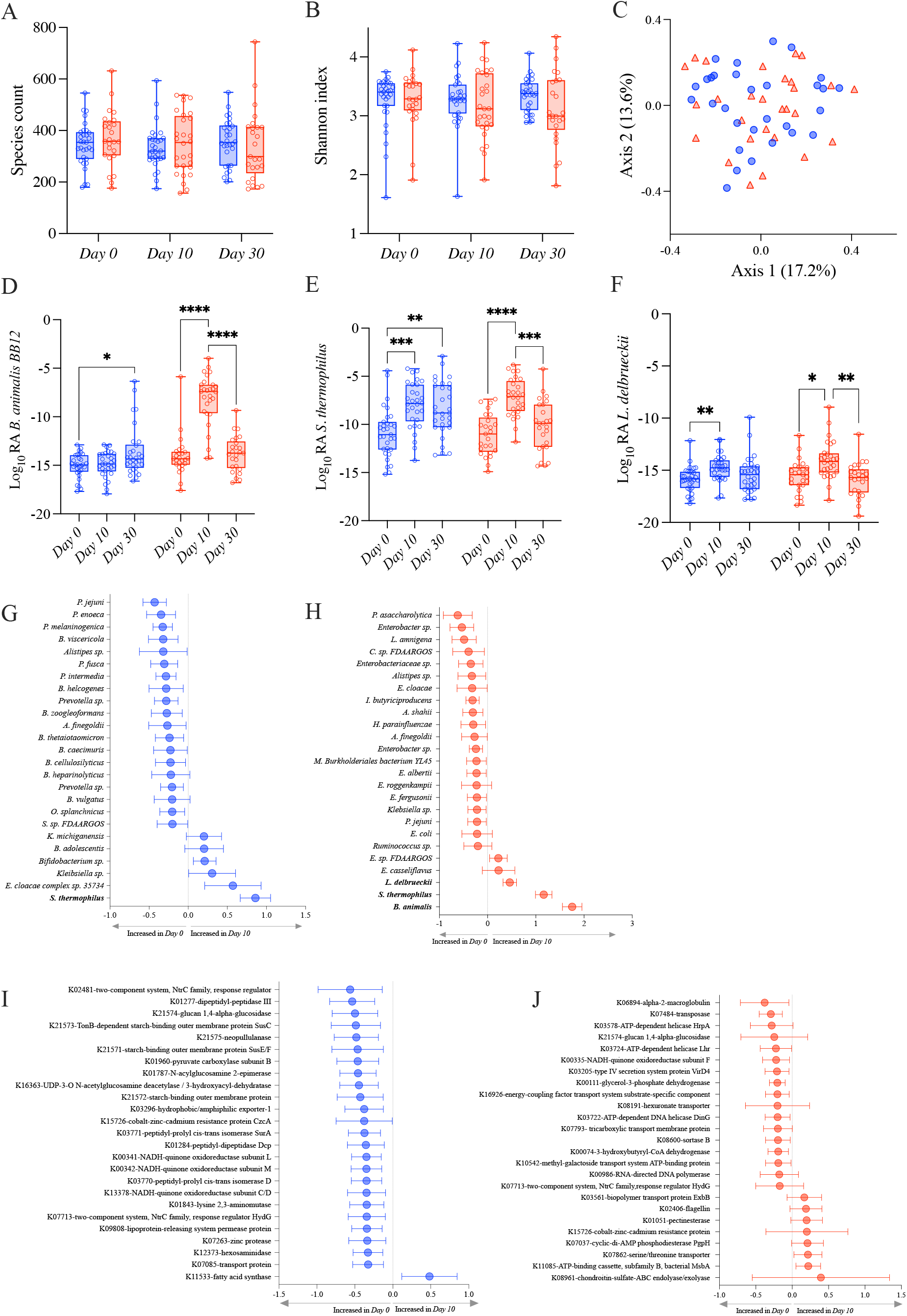
Observed species (**A**) and Shannon (**B**) alpha diversity indices showing no differences comparing S2 (blue) and S2+BB12 (red) groups. PCoA based on Bray-Curtis dissimilarity followed by a PERMANOVA (*p*=.318) showing no separation among samples. Relative abundances of the three strains used in the S2 and S2+BB12 interventions: *B animalis* (**D**), *S. thermophilus* (**E**), and *L. delbruekii* (F) during time comparing S2 (blue) and S2+BB12 groups (red). Top 25 changing taxa (**G-H**) and gene orthologs (**I-J**) between *Day 0* and *Day 10* in S2 (**G-I**) and S2+BB12 (**H-J**) groups identified by linear model.

We built upon these findings by using shotgun metagenomics, which allowed us to observe differential abundant taxa at the species level. As expected, the metagenomic analysis showed an increase of *B. animalis* in the S2+BB12 group. *B. animalis* abundance was significantly impacted by the administration of the BB12 supplemented yogurt (group × time *p* < .001; Figure 2D). We observed an effect of time, but not group, in *L. delbrueckii* (time *p* <.001; group *p* = .405). *S. thermophilus*’s relative abundance were significantly different in time and group x time interaction (time *p* <.001; group *p* = .349; group x time *p* = .020) (Figure 2E-F). As expected, *post hoc* testing showed in the S2+BB12 group a statistically significant increase comparing *Day 0 vs. Day 10* in BB-12 (*p* < .001), *S. thermophilus* (*p* < .001), and *L. delbrueckii* (*p* = .018) abundances. Similarly, *Day 10 vs. Day 30* comparison showed a statistically significant decrease in BB-12 (*p* < .001), *S. thermophilus* (*p* = .001), and *L. delbrueckii* (*p* = .004) abundances (Figure 2D-F). In the S2 group, we found statistically significant differences in *Day 0 vs. Day 10* comparison of *S. thermophilus* (*p* < .001) and *L. delbrueckii* (*p* = .005) abundances. Interestingly, BB-12 increased comparing *Day 0* to *Day 30* in the same group (*p* = .016).

We then focused our analyses on within-group differences in both groups from *Day 0* to *Day 10*, as both groups received a probiotic intervention (S2 *vs*. S2+BB12) and were healthy children. The 25 taxa with the greatest change in relative abundance between *Day 0* and *Day 10* were evaluated in order to observe the effect on individual microbial taxa following a ten-day administration of two (S2) *vs*. three (S2+BB12) strains of probiotics. In addition to the intervention bacteria that changed in the S2+BB12 group, we observed that the mean relative abundance was lower than the control group for seven taxa belonging to *Prevotella* genus, though the difference was not statistically significant after correction for multiple comparisons: *P. intermedia* (*p* = .999), *P. melaninogenica* (*p* = .574), and *P. jejuni* (*p* = .444) (Figure 2G, Supplemental Table 1). Other top changing bacteria S2+BB12 group were decreased relative abundance in bacterial species from *Day 0* to *Day 10*, but they did not remain statistically significant after FDR correction including *I. butyriciproducens* (*p* = .999), *Enterobacter sp. FY-07* (*p* = .999), and *P. asaccharolytica* (*p* = .999; Figure 2H, Supplemental Table 1).

When we analyzed the differential abundance of gene orthologs, we found that the relative abundance of the glucan 1,4-alpha-glucosidase and response regulator HydG genes decreased from *Day 0* to *Day 10* in both groups, (Figure 2I-J). These differences were also not statistically significant before or after FDR correction (Supplemental Table 1). Except for one ortholog (fatty acid synthase [K11533]), all the 25 top changing gene orthologs were decreased in the S2 group after ten days of probiotics administration, although these decreases were not statistically significant (Figure 2I). An overall trend of decreased orthologs relative abundance from *Day 0* to *Day 10* was similar in the S2+BB12 group, except for eight orthologs that had a non-statistically significant increase following BB12 administration (Figure 2J).

We selected 18 known probiotic strains identified through literature search (Supplemental Table 2), to observe if the administration of two (S2) *vs*. three (S2+BB12) probiotic strains would influence the abundance other known probiotics. Most of the identified probiotic-associated taxa belonged to *Lactobacillus, Bacteroides*, and *Bifidobacterium* genera, and were commonly used as probiotic supplements in the food industry or clinical trials (see Supplemental Table 2 for references). We observed that the abundance of targeted probiotic-associated bacteria increased from *Day 0* to *Day 10* in 61% (11/18) of selected taxa in the S2 group (Figure 3A) and 72% (13/18) of taxa in the S2+BB12 (Figure 3B) group, but most of the differences did not reach statistical significance. Nevertheless, cumulative bacterial responses changes of probiotic-associated bacteria differed according to intervention group. For example, we observed no pattern in samples distribution in the S2 group (Figure 3C), while in the S2+BB12 group we can observe a separation of *Day 0 vs. Day 10* samples (Figure 3D). In the S2+BB12 group, many of the *Day 10* probiotic-associated bacteria clustered with *B. animalis*, indicating similar response to the probiotic (Figure 3D).

**Figure 3:**
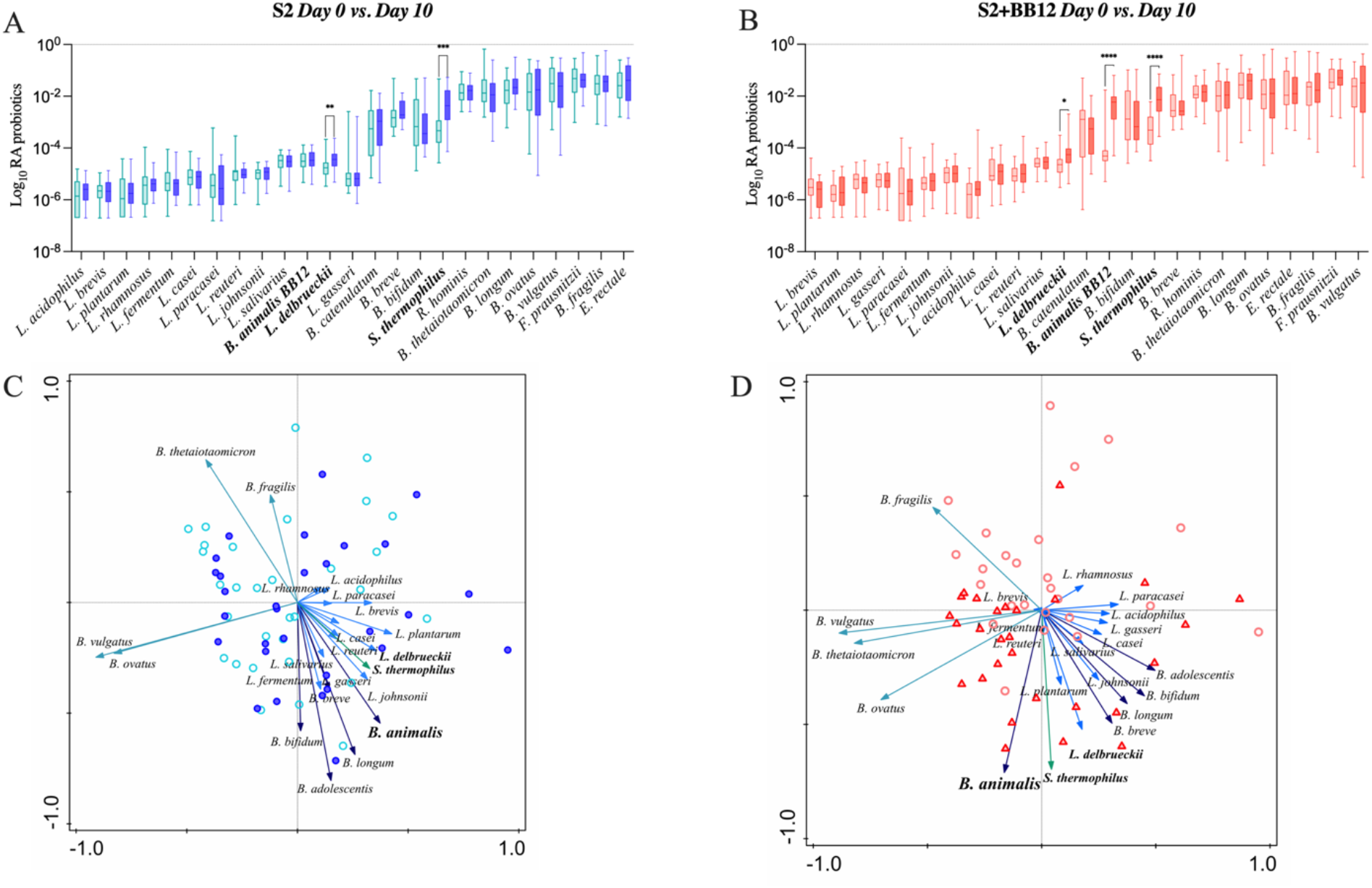
Relative abundances of selected probiotic taxa are plotted in S2 (**A**) and S2+BB12 (**B**) groups after ten days of probiotic intervention (*Day 0 vs. Day 10*). Post-selection PCA shows selected probiotics following ten days of probiotics administration in S2 (**C**) and S2+BB12 (**D**) groups. The difference in *S. thermophilus* and *B. animalis* between *Day 0* and *Day 10* abundances are statistically significant different (*p* < .0001 for both) in S2+BB12 group. *Day 10* is represented by the darkest color all plots. In PCA, different arrows colors refer to different genera: Blue, light blue, dark green, and green represent *Bifidobacterium, Lactobacillus, Bacteroides*, and *Streptococcus* genera respectively. Each arrow points in the direction of the steepest increase of the values for the corresponding feature. The angle between arrows indicates the correlation between the different features (positive when the angle is sharp and negative when the angle is larger than 90 degrees). The length of the arrow is a measure of fit for the feature.

### Metabolomic analyses

The untargeted metabolomic analysis identified 734 metabolites. To evaluate the impact of S2 *vs*. S2+BB12 probiotics on the metabolome, we performed both exploratory analyses quantifying differences in all annotated metabolites and hypothesis-driven metabolomics analyses focused on amino acid-associated and short chain fatty acid metabolites.

After excluding xenobiotics, we conducted exploratory metabolomic analyses on 601 biochemicals. We analyzed the differences between *Day 0 vs. Day 10, Day 10 vs. Day 30, and Day 0 vs. Day 30* within the S2 and S2+BB12 groups (Supplemental Table 3). Metabolites changes [t-test (raw *p*-value=.05) and FC analysis (threshold = 2)] comparing *Day 0 vs. Day 10* and *Day 0 vs. Day 30* in S2 (Figure 4A, C, E) and S2+BB12 (Figure 4B, D, F) groups are presented. We found greater changes in the S2+BB12 group compared to the S2 group (14 *vs*. three metabolites). After ten days of yogurt consumption, we found an increase of N-acetylvaline (raw *p*=.022 FC=2.151) and a decrease of arachidoylcarnitine (C20) * (raw *p*=.045 FC=.436) and nicotinate ribonucleoside (raw *p=*.009 FC=.326). *Day 0 vs. Day 30* comparison exhibited several changes in lipids decreased at Day 30 belonging to diacyglicerol metabolism: linoleoyl-linoleoyl-glycerol (18:2/18:2) [1]* (raw *p*=.016 FC=0.29), palmitoyl-linoleoyl-glycerol (16:0/18:2) [2]* (raw *p=*.009 FC=.311), palmitoyl-linoleoyl-glycerol (16:0/18:2) [1]* (raw *p*=.008 FC=.263), oleoyl-linoleoyl-glycerol (18:1/18:2) [2] (raw *p=*.007 FC=.295), palmitoyl-oleoyl-glycerol (16:0/18:1) [2]* (raw *p*=.004 FC=.252), and oleoyl-linoleoyl-glycerol (18:1/18:2) [1] (raw *p*=.002 FC=0.221). The glycerolipids 2-palmitoyl-galactosylglycerol (16:0)* (raw *p*<.001 FC=2.358) and 1-palmitoyl-galactosylglycerol (16:0)* (raw *p=*.001 FC=2.319), were increased together with 3-hydroxybutyrate (BHBA) (raw *p=*.022 FC=2.821). Comparing *Day 10 vs. Day 30*, we found adenine decreased at *Day 30* (raw *p=*.023 FC=0.48).

**Figure 4:**
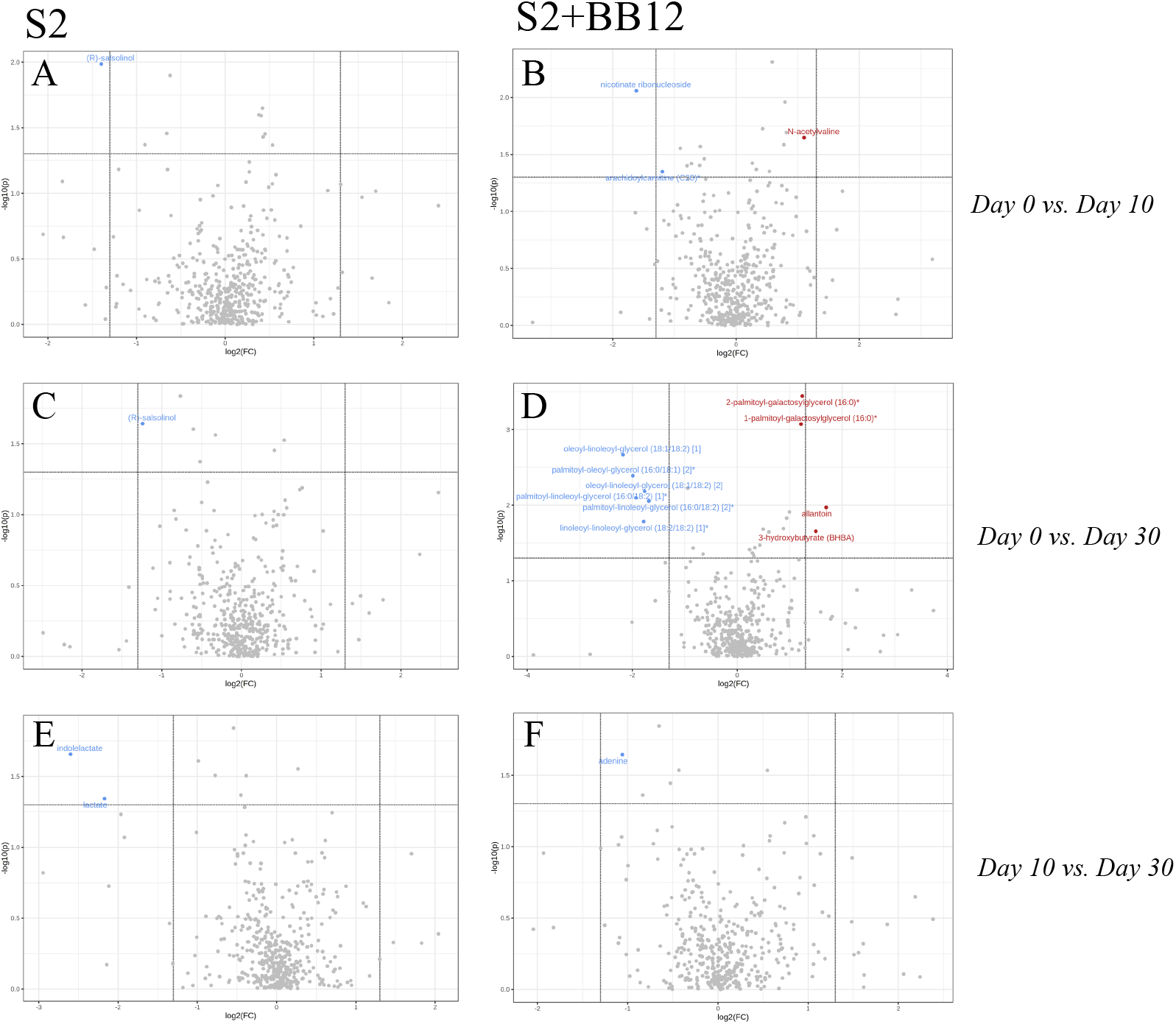
Volcano plot (*p* < .05, FC>2) comparing *Day 0 vs. Day 10* (**A, B**), *Day 0 vs. Day 30* (**C, D**), and *Day 10 vs. Day 30* **(E, F**) within S2 and S2+BB12 respectively. Metabolites in red/blue represent metabolites increased/decreased at *Day 10* (**A, B**) and *Day 30* (**C-F**).

In the S2 group, the amino acid (R)-salsolinol was decreased after ten days of intervention (*Day 0 vs. Day 10*) (raw *p=*.01 FC=.38) and 20 days after yogurt consumption discontinuation (*Day 0 vs. Day 30*) (raw *p=*.023 FC=.423). Lactate and indolelactate were also decreased looking at *Day 0 vs. Day 30* comparison (FC=0.222, raw *p=*.045 and FC=0.165, raw *p=*.022, respectively). PCA of untargeted metabolites showed no separation between the time points in any group (Supplemental Figure S2 A-F).

Next, we focused on evaluating differences in specific metabolites associated with amino acid metabolism or biosynthesis, as we hypothesized that they would be impacted by probiotic intervention and associated with the gut microbiome in both groups. There were several amino-acid associated metabolites that differed as a result of time in both groups (Supplemental Table 4) including 3-methyl-2-oxobutyrate (*p*=.004), 3-methyl-2-oxovalerate (*p*=.025), alanine (*p*=.001), glutamate (*p*=.034), isoleucine (*p*=.020), and valine (*p*=.004), among others. Conversely, glycine (*p*=.025), indolelactate (*p*=.034), N-acetylserine (*p*=.049), and pyroglutamine (*p*=.007) had significant group x time effects as a result of the S2 and S2+BB12 probiotic interactions (Supplemental Table 4). In the S2+BB12 group, average fecal metabolite abundances of 4-methyl-2-oxopentanoate, alanine, cysteine s-sulfate, glycine, histidine, lysine, N-acetylalanine, N-acetylglutamine, N-acetylleucine, phenylalanine, serine and valine were significantly lower at *Day 10* (versus baseline *Day 0;* Figure 5 A-N). Alanine, glycine, and N-acetylglutamine metabolite levels continued to be lower twenty days after the S2+BB12 probiotic was discontinued (*Day 30*) versus baseline (Figure 5B, D, H), while fecal cysteine s-sulfate levels increased back to baseline levels after probiotic discontinuation.

**Figure 5.**
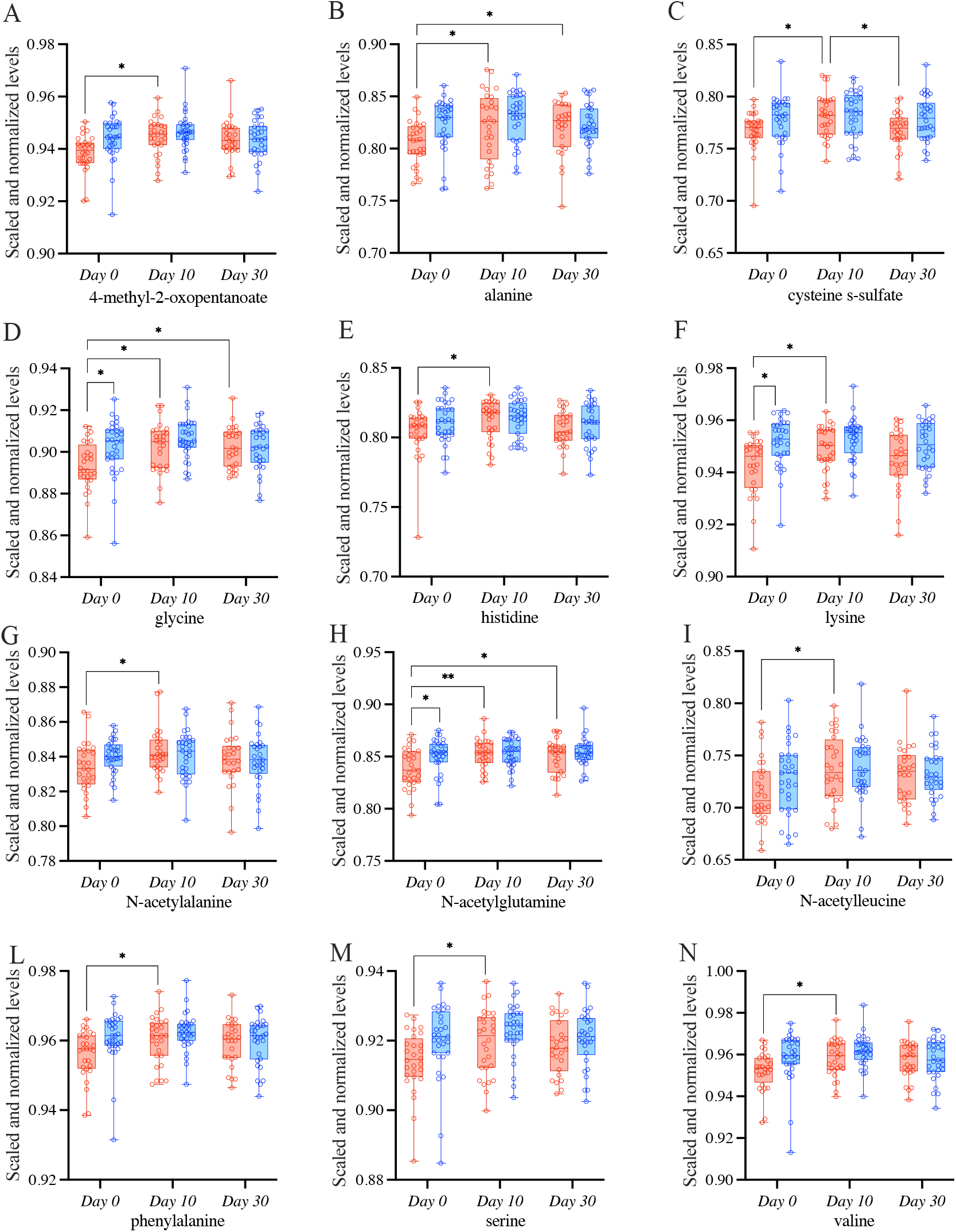
Targeted metabolomic analysis on amino acids was used to study the differences comparing *Day 0 vs. Day 10, Day 0 vs. Day 30*, and *Day 10 vs. Day 30* within S2 (blue) and S2+BB12 (red) groups. Linear mixed-effects model followed by post hoc pairwise testing with Tukey’s correction (when appropriate) were performed. All mixed model results of selected amino acid-associated metabolites and post hoc testing results (when group, time, or group * time model results were significant) are listed in Supplemental Table 3.

### Integration of Metagenomics and Metabolomics Datasets

Microbe-metabolite interactions were tested through correlation matrices and visualized with network graphs. After filtering, 26 microbial taxa and 79 metabolites were tested for associations (Figure 6, Supplemental Table 5-6).

**Figure 6:**
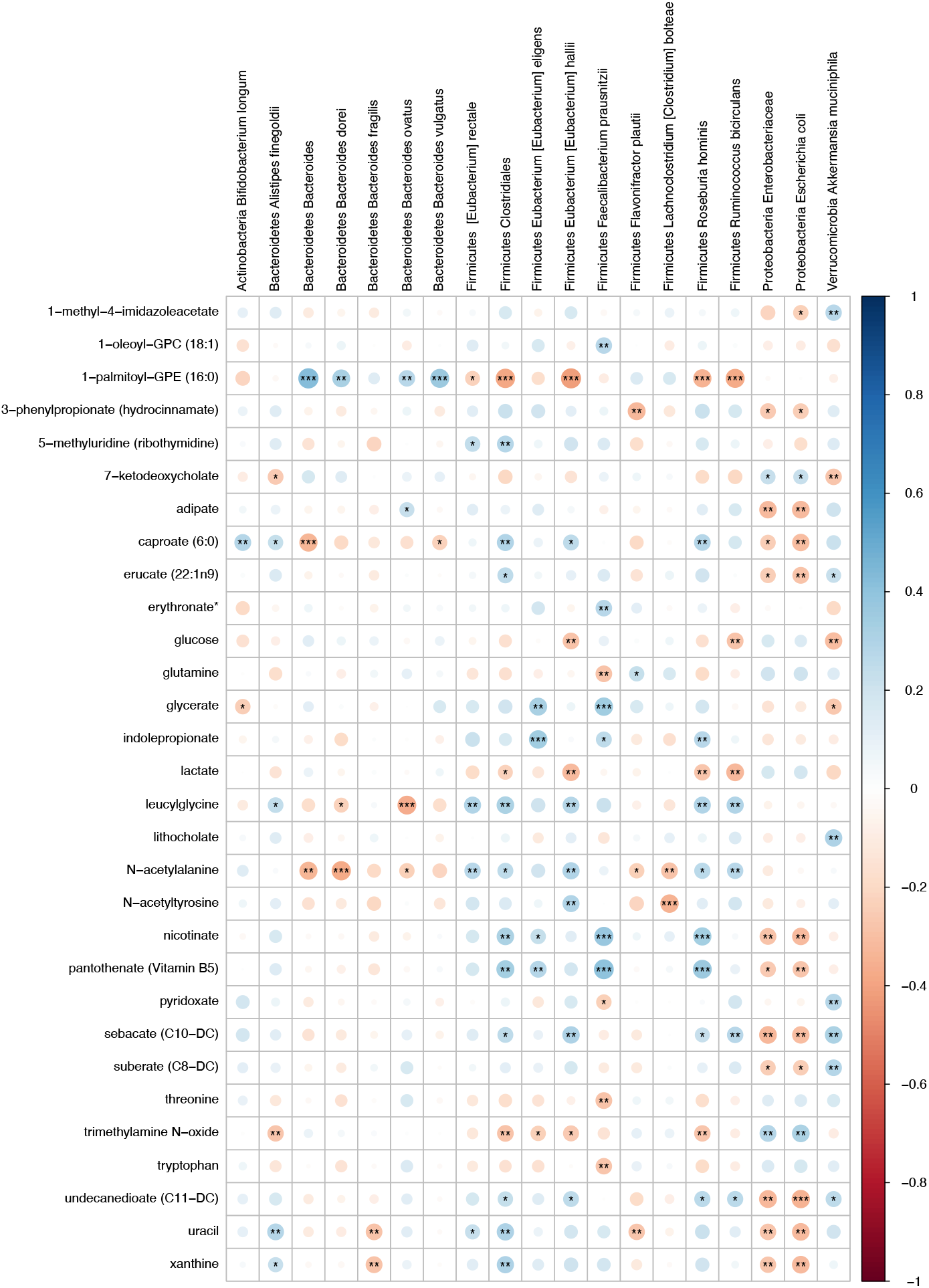
Heatmap representing FDR corrected correlations between microbial taxa and metabolites in S2 and S2+BB12 groups. Red and blue represent negative and positive correlations respectively. Stars indicate FDR values: * = FDR < .05, ** = FDR < .01, and *** = FDR < .001.

Metabolites that were significantly positively correlated with Enterobacteriaceae and *Escherichia coli* included trimethylamine N-oxide (TMAO), ursodeoxycholate, 7-ketodeoxycholate, and glycine (Figure 7). Other taxa that had multiple significant positive associations with metabolites included *Akkermansia muciniphila, Eubacterium hallii, Roseburia hominis*, and Clostridiales. Of the supplemented probiotic bacteria, *B. animalis* was positively associated with uracil, *S. thermophilus* with deoxycarnitine and phenylalanine, and *L. delbrueckii* with thymine. Known products of bacteria such as nicotinate (vitamin B3), pantothenate (vitamin B5) correlated positively with Clostridiales, *R. hominis, F. prausnitzii*, and *E. eligens* (Figure 7A).

**Figure 7.**
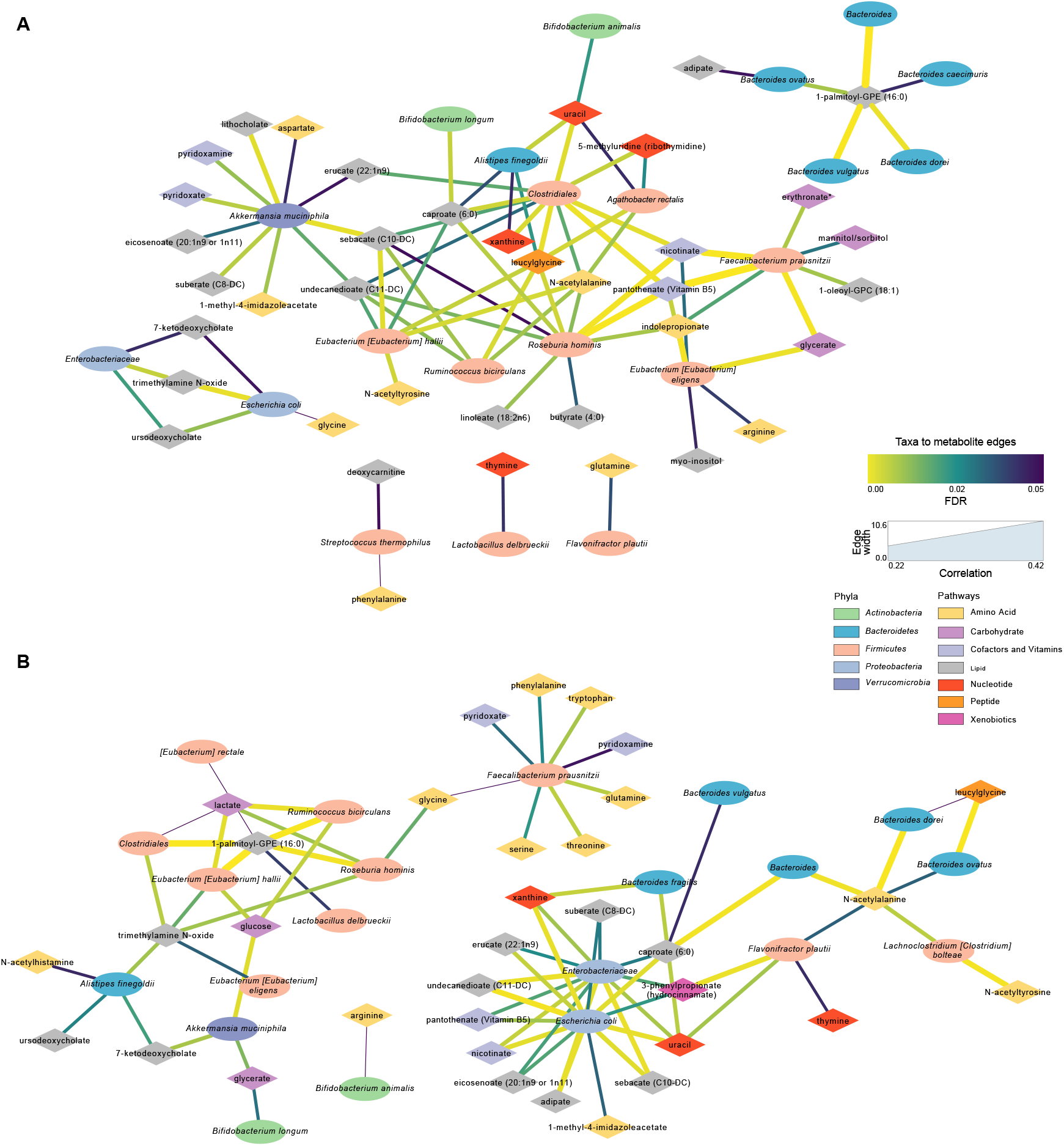
Significant microbial taxa and metabolite associations: Network analyses in samples across *Day 0, Day 10*, and *Day 30* within S2 and S2+BB12 groups. **A**. Network generated from microbial taxa (ellipses) that passed filtering and microbial-associated metabolites (diamonds) identified from previous work [38] showing only positive correlations. **B**. Same network but showing only negative correlations. Width of edges is proportional to Spearman correlation coefficient and edge color is mapped to the FDR value or the correlation test. All correlations between selected taxa and metabolites and statistical significance testing results are listed in **Supplemental Table 5**.

Several amino acids correlated negatively with *F. prausnitzii* and *R. hominis*, including glutamine, lactate, threonine, tryptophan and TMAO (Figure 7B). Lactate was also negatively associated with *A. rectalis, R. bicirculans, E. hallii*, and Clostridiales; glucose with *R. bicirculans, E. hallii*, and *A. muciniphila*; and glycerate with *A. muciniphila* and *B. longum*. Among the probiotics of interest, *B. animalis* correlated negatively with arginine and *L. delbrueckii* with 1-palmitoyl-GPE (16:0). Uracil was positively associated with Clostridiales, *A. finegoldii*, and *E. rectale*. Uracil was also negatively correlated with *B. dorei, F. plautii*, Enterobacteriaceae and *E. coli*.

## DISCUSSION

This randomized, controlled study aimed to observe the differences following the administration of yogurt supplemented with two (S2) *vs*. three (S2+BB12) probiotic strains in healthy children. We studied probiotics consumption effects on the fecal microbiome and metabolome comparing baseline, post-ten days (*Day 10*), and post-30 days (*Day 30*) following yogurt administration. There were no significant differences in the global metagenomic or metabolomic profiles between healthy children receiving two (S2) *vs*. three (S2+BB12) probiotic strains for 10 days. Nevertheless, we observed a significant increase (*Day 0* to *Day 10*) in the relative abundance of the two and three probiotics administered in the S2 and S2+BB12 groups, respectively, indicating the intervention had a measurable impact on the bacteria of interest in the gut microbiome. Interestingly, in the S2 group, *S. thermophilus* species appear to maintain a sustained increase over time, maintaining the significant increase in relative abundance from *Day 0* vs. *Day 30* As both cohorts of children received a probiotic cocktail and the intervention lasted over a short period of time, the lack of strong association between groups in global microbiome changes is not surprising and suggests a resilient and stable gut microbiota in this cohort of healthy children ages one to five. Nevertheless, existing literature reports about infant’s and children’s gut microbiota stability are conflicting. [39] [2] Infant and children’s gut microbiota are thought to be immature and therefore more susceptible to perturbations. In line with other studies [40, 41] our data show short-term effects of probiotic administrations in both groups on the gut microbiome and metabolome. The children’s gut microbiota stability observed in this study is consistent with the parent study’s findings. [30]

Most studies examining the health benefits of probiotics have focused on people with pathologies. However, few studies have examined the effects of probiotics on healthy individuals and even fewer have examined the use of probiotics in children. A review of probiotic supplementation in healthy adults found that probiotic supplementation led to a transient increase in the concentration of supplement-specific bacteria but failed to support the ability of probiotics to cause persistent changes in gut microbiota. [42] This is consistent with our results finding a significant increase in the concentration of the supplement-specific bacteria BB-12, following ten days of BB-12 supplementation. Importantly, as reported in adults, these results were temporary. On *Day 30* (20 days following the termination of the BB-12 supplementation), there were no significant differences in the concentration of BB-12 in the S2+BB12 group.

As yogurts and other dairy products commonly supplemented with probiotics often possess other beneficial characteristics, such as a high calcium content, these properties of probiotics could allow consumers to benefit from the nutritional components without risking disruption to their microbiota and health. This potential beneficial shift to probiotic-associated bacteria that we demonstrated in the S2+BB12 group (Figure 3) supports the theory that a probiotic intervention may provide a net positive contribution to the gut microbiome ecosystem without conferring strong effects on specific individual bacteria. These findings are also consistent with the concept of emergent properties which postulates that individual properties cannot entirely be explained by their individual components. [43] Therefore, in the context of the current study, although no significant differences following the interventions were observed, there was a global shift towards the increase of bacteria belonging to *Bifidobacterium* and *Lactobacillus* genera, commonly used probiotics. The combined effects of these bacteria could have more functional implications and protective effects than individual bacterial changes supporting the use of probiotics to prevent gastrointestinal disorders in children, although more research is needed to validate this notion.

Species belonging to *Bifidobacterium* genus were highly abundant in both groups at the three time points. Bifidobacteria are highly represented in children, as they use milk oligosaccharides as a carbon source and to restrict human milk oligosaccharides availability to other microorganisms [44]. In addition to the parent study findings, our metagenomic approach allowed a low-level taxonomic affiliation, revealing differences among species belonging to the same genus as in the case of *Bifidobacterium*. It explains why *Bifidobacterium* genus was not among the differentially abundant species in the previous study, *i*.*e*., because different *Bifidobacterium* species were abundant in the experimental group (S2+BB12) and in the control group S2 (*e*.*g*., *B. catenulatum, B. pseudocatenulatum*). Additionally, this metagenomic approach adds to the existing literature of the genomic potential of the microbial community underlying microbiome-host interactions.

Because the fecal metabolome is influenced by different factors, these changes could reflect shifts of dietary intake, digestion, microbial degradation, and host absorption. When we analyzed the fecal metabolome and microbial diversity using both participant groups, we found associations between metabolites (*e*.*g*., uracil, deoxycarnitine, and thymine) and microorganisms. Uracil was positively associated with Clostridiales family, *A. finegoldii, B. animalis*, and *E. rectale. B. dorei, F. plautii*, Enterobacteriaceae family, and *E. coli* were negatively correlated with uracil. During infections, the immune response triggered by uracil promotes pathogen bacteria elimination, intestinal cell repair, and host homeostasis. [45] Thymine was positively associated with *L. delbrueckii*, the probiotic used in both interventions, and negatively correlated to *F. plautii*. Thymine was found to accelerate microbial metabolisms and ROS production improving antibiotic efficacy both *in vitro* and *in vivo*. [46] Deoxycarnitine, positively associated with *S. thermophilus*, was linked to increased intestinal permeability. [47] This suggests that the fecal metabolome may influence gut immune function, permeability, and homeostasis.

A strength of our study is that it was a randomized, blinded controlled trial conducted on children ages one to five years old, an age group that is rarely studied in probiotic research. Additionally, we incorporated and integrated both metagenomic and metabolomic analyses to characterize the effect of BB-12 supplemented yogurt on the children’s gut-microbiome and metabolism. This unique integration allowed us to test more system-wide and functional effects on the gut microbiome as a result of two probiotic interventions, providing more comprehensive data on gut health. However, this study examined short-term changes of probiotics over a ten-day period, and more studies should be conducted to investigate the long-term effects on probiotic consumption in this age group. Additionally, we did not incorporate information of dietary intake in the current analysis.

More studies are needed to elucidate the mechanistic pathways by which probiotics such as BB-12 can affect mucosal barrier functions and innate immunity. Future studies should expand upon the findings presented in this double-blinded, randomized controlled trial and examine the interplay of diet and probiotics on metabolites and microbiota in children. Additionally, further research is needed to investigate environmental factors that influence the impacts of probiotics on children’s health status and behavior. The long-term supplementation BB-12 on this population and its longitudinal effects during development should also be examined. This would allow for more in-depth knowledge of the impact that probiotics have on gut microbial communities in developing children as they age. Additionally, future research in children at risk for gastrointestinal disorders may elucidate if these functional metabolite changes as a result of S2+BB12 probiotic intervention confer a protective gastrointestinal effect.

In conclusion, the results from this deep metagenomic and metabolomic characterization of the gut microbiome and metabolome of children following BB-12 consumption did not show significant differences between the groups, although net positive emergent property effects were witnessed in the S2+BB12 group over time. The functional redundancy in healthy microbial systems and metabolic stability reflects no changes in the microbial diversity, although we did observe a separation effect in the S2+BB12 group as a result of the three-strain intervention when we focused on probiotic-associated and beneficial bacteria reported the literature. Our study validated previous results from Tan *et. al* and allowed a more in-depth taxonomic characterization of the microorganisms, their genes, and their metabolites. We detected higher abundances of two of the probiotic intervention bacterial taxa (*B. animalis* and *S. thermophilus*) in study subjects receiving the based S2 probiotic intervention + BB12, but no individual taxonomic changes occurred in the S2 only group. Finally, although we did not see global fecal metabolome response to either probiotic, several fecal metabolites were decreased in the S2+BB12 group, indicating a net functional impact of the addition of BB12 to the probiotic intervention. Future research replicating these results across different patient populations will confirm the therapeutic use of BB12 as a probiotic intervention to exert beneficial impacts on the pediatric gastrointestinal system.

## Materials and Methods

### Study Design, Participants, and Setting

Participants ages one to five years old were recruited through the Capital Area Primary Care Research Network for phase I of a double-blinded, randomized controlled study (protocol NCT001652287). Participants included in this study were children whose parents/caregivers were able to read, write, and speak either English or Spanish and had access to a telephone and refrigerator. Eligible participants provided written informed consent were enrolled and randomized as described by Tina et al to either the BB-12^®^ or control yogurt drink by family cluster. The study protocol was approved by the Georgetown University Institutional Review Board (IRB No. 2012-1112, Washington, DC). The independent Data Safety Monitoring Board reviewed the protocol before study initiation and checked adverse event data at approximately 33%, 50% and 66% data completion. Additional monitoring was conducted by the FDA/CBER, under IND#13691 and the National Institutes of Health (NIH), National Center for Complementary and Integrative Health (NCCIH), including its Office of Clinical and Regulatory Affairs. Participants’ eligibility criteria are described in Tan et al., [30] which included the absence of lactose intolerance and chronic conditions, such as diabetes and asthma. The participants were asked not to consume any products containing probiotics for 14 days before initiating the yogurt intervention and throughout the entire intervention period. The base yogurt was prepared with live starter cultures of *Streptococcus thermophilus* and *Lactobacillus delbrueckii* probiotics (referred to as the two strain [S2] yogurt group), as described in Tan et al. [30]. At baseline, the children were randomized into two groups called S2 or S2+BB12. Participants in the S2 group (n = 31) were administered 112 g of the base yogurt beverage (containing *Streptococcus thermophilus* and *Lactobacillus delbrueckii* only). In contrast, participants in the S2+BB12 group (n = 28) were administered the base yogurt beverage, with an additional 1×10^10^ colony-forming units of BB12 per serving per day. The yogurt was administered to the participants for ten consecutive days.

#### Sample Collection and Processing

Fecal samples were collected at baseline (*Day 0*), following ten days of yogurt consumption (*Day 10*), and 20 days following discontinuation of yogurt administration (*Day 30*), and immediately stored after collection at -80 ° C. Samples from days zero, ten and 30 were then thawed, and approximately 100 mg of the samples were sent to Microbiome Center of the Children’s Hospital of Philadelphia (n = 169) for microbiome analysis and another 100mg were sent to Metabolon Inc. (Morrisville, NC, USA; n = 174) for metabolomic analyses.

### Metagenomic profiling

The DNA used for the metagenomic analysis was extracted using the DNeasy PowerSoil Kit (Qiagen, Hilden, Germany) and quantified with the Quant-iT PicoGreen Assay Kit (Molecular Probes). Shotgun libraries were generated from 0.5ng DNA using the Nextera XT Library Prep Kit (Illumina, San Diego, CA, USA) and libraries were sequenced on an Illumina HiSeq 2500 in High Output mode to produce paired-end 125bp sequence reads. Extraction blanks and nucleic acid-free water were processed along with experimental samples to empirically assess environmental and reagent contamination. A laboratory-generated mock community consisting of DNA from *Vibrio campbellii* and Lambda phage was included as a positive sequencing control.

### Metabolomic analysis

The metabolomic analysis was performed using untargeted ultra-performance liquid chromatography-tandem mass spectrometry (UPLC/MS/MS, Waters ACQUITY, Milford, MA, USA), as described previously [48]. Briefly, the fecal samples were prepared using the automated MicroLab STAR system (Hamilton Company, Franklin, MA, USA) and extracted at a constant per-mass basis. Proteins were removed using methanol precipitation (Glen Mills GenoGrinder 2000), followed by centrifugation. The samples were processed using four methods: reverse phase (RP)-UPLC/MS/MS with electrospray ionization (ESI), in both positive (optimized for hydrophilic and hydrophobic compounds, respectively) and negative modes, and hydrophilic interaction chromatography (HILIC)-UPLC/MS/MS-ESI in negative ion mode. The raw UPLC/MS/MS data were integrated into ion peaks organized by mass, retention time/index, and peak area. Metabolites were annotated by comparison of individual spectra to a standard reference library, and area-under-the-curve analysis was performed for peak quantification.

### Statistical Analysis

For gut microbiome and metabolome analyses, we studied within and between group differences after ten days of probiotic consumption (*Day 10*) and 20 days post probiotic discontinuation (*Day 30*). Shotgun metagenomic data were analyzed using Sunbeam. [49] The abundance of bacteria was estimated using Kraken. [50] Taxa that were above 0.1% abundance in any sample were used for differential abundance testing, and differential abundance analysis was performed using linear models of Log_10_ transformed relative abundances. Reads were mapped to the KEGG database [51] using Diamond [52] to estimate the abundance of bacterial gene orthologs. Alpha diversity within samples in the S2 and S2+BB12 groups were assessed by computing the expected number of species at a sequencing depth of 1,000 reads and the Shannon index. To evaluate community-level differences between S2 and S2+BB12 group fecal samples, beta diversity was calculated using Bray-Curtis dissimilarity matrices, visualized using Principal Coordinates Analysis (PCoA) plots, and relationships within and between S2 and S2+BB12 groups were compared using the PERMANOVA test.

The top 25 most abundant bacterial taxa and gene orthologs from shotgun metagenomics sequencing were selected using linear models to evaluate the taxa with greatest estimated change in Log_10_ transformed relative abundance for the given comparison. Probiotic-associated bacteria, previously demonstrated to be short chain fatty acid (SCFA)-producers and beneficial for GI health, [53] were determined from the literature (Supplemental Table 2). The impact of the S2 *vs*. S2+BB12 probiotic strains on overall probiotic-associated bacterial relative abundance was additionally measured and visualized by CANOCO version 5 [54] in a post-selection PCA to evaluate the effect of targeted probiotic strain administration on bacterial responses of taxa known to be linked to gut microbiome health.

Exploratory and hypothesis-driven metabolite analyses were performed with untargeted metabolite data processed by Metabolon Inc. using MetaboAnalyst 5.0 (https://www.metaboanalyst.ca/)[55] and R. [56] Metabolites with 20% or more missing values were excluded from the exploratory analyses. Missing values, if any, were imputed as 1/5 of the minimum positive value of each feature. Metabolite values were median-scaled and Log_10_ transformed. Wilcoxon rank-sum test, fold change (FC) analyses (FC threshold = 2), and Principal Component Analysis (PCA) were performed to analyze differences between *Day 0 vs. Day 10, Day 0 vs. Day 30, and Day 0 vs. Day 30* within the S2 and S2+BB12 groups and between groups within each time point.

Hypothesis-driven metabolite analyses were additionally performed in metabolites associated with the amino acid super pathways (*i*.*e*., glycine, serine and threonine metabolism, alanine and aspartate metabolism, *etc*.), and SCFAs, as these metabolites are strongly associated with gut microbial community characteristics. [9] Linear mixed-effects model followed by *post hoc* pairwise testing (when appropriate) and Tukey’s correction were performed in JMP statistical analysis platform. [57]

To create a network of metabolite-taxa correlation pairs, filtering was applied to metabolites as above as well as restricting to bacterial substrates and products based on previous work [58]. Briefly, metabolite substrates were defined as those that were increased after treatment with antibiotics and products were those that decreased. [38] Microbial taxa at the species level were filtered to include taxa present at > 0.01% mean relative abundance and taxa that changed in the S2+BB12 treatment group (*i*.*e*., *Lactobacillus delbrueckii, Streptococcus thermophilus*, and BB-12). Spearman correlation testing was then performed on each microbe-metabolite pair with FDR correction applied to p-values. Network diagrams of bacteria and metabolites were generated using Cytoscape v3.9.1. [59] Metabolites that significantly correlated with *L. delbrueckii, S. thermophilus*, and BB-12 were additionally tested for intervention-associated change over time. Statistical significance was defined as *p*-values or FDR corrected *p*-values < .05 for all statistical analyses.

## Data Availability

All data produced in the present study are available upon reasonable request to the authors. Data was also submitted to SRA under project number PRJNA929986.

https://dataview.ncbi.nlm.nih.gov/object/PRJNA929986

## Abbreviations

FC: Fold Change
FDR: False Discovery Rate
GI: Gastrointestinal
PCA: Principal Component Analysis
PCoA: Principal Coordinate Analysis
PERMANOVA: Permutational Analysis of Variance
RA: Relative Abundance
S2: Two-strain (*Streptococcus thermophilus* and *Lactobacillus delbrueckii*) Yogurt Group
S2+BB12: Two-strain (*Streptococcus thermophilus* and *Lactobacillus delbrueckii*) plus *Bifidobacterium animalis* subsp. *lactis* strain BB-12 Yogurt Group
TMAO: Trimethylamine N-oxide

## Figures and Tables

Figure 1AB = Study Design and Intervention

Figure 2AJ = Metagenomic results

Figure 3AD = Other Probiotics Comparison

Figure 4 AF = Metabolite Volcano Plots

Figure 5AN = Selected Metabolites Analysis

Figure 6 = Heatmap Microbiome/Metabolome

Figure 7AB = Network Analysis (Metagenomics/Metabolomics)

Figure S1 = Metagenomics PCoA Day 30

Figure S2 = Metabolite PCA

Table 1 = Participants Demographics

Supplemental table 1 = Top_25_linear_model_KEGG

Supplemental table 2 = Probiotics selection

Supplemental table 3 = Metabolites and Pathways Volcano plot

Supplemental table 4 = Selected_metabolite_model_results

Supplemental table 5 = Table_S5_26_taxa_to_479_metab_correlations

Supplemental table 6 = Table_S6_26_taxa_to_79_metab_network

## Acknowledgements

The authors would like to thank Dr. Joan Austin for her review and edits and Dr. Jennifer J. Barb for bioinformatics consultative support. A special thanks to the participants of this study and the Georgetown Microbiome Group.

## Author contributions

Concept and design: Drs. Merenstein and Joseph

Acquisition of data, sample processing: Tan, Dr. Merenstein, Ms. Franks, Ms. Brooks

Statistical Analysis, or interpretation of data: Dr. Vizioli, Ms. Franks, Dr. Daniel, Dr. Bittinger, Dr. Jaime-Lara, Dr. Maki, Dr. Joseph

Drafting of the manuscript: Dr. Vizioli, Dr. Diallo, Ms. Franks, Ms. Brooks, Dr. Jaime-Lara, Dr. Maki, Dr. Joseph

Critical revision of the manuscript for important intellectual content: Dr. Diallo, Dr. Merenstein, Dr. Tan, Brooks, Dr. Joseph, Dr. Scott, Dr. Bittinger

Administrative, technical, or material support: Dr. Joseph

Obtained funding: Drs. Joseph & Merenstein

Study supervision: Drs. Merenstein & Joseph

## Author’s Note

The content is solely the responsibility of the authors and does not necessarily represent the official views of the National Institutes of Health.

