## Supplemental Table 2 for "Administration of *Bifidobacterium animalis* subsp. *lactis* Strain BB-12^®^ in Healthy Children: Characterization, Functional Composition, and Metabolism of the Gut Microbiome"

Supplemental Table 1. Probiotic Associated Bacteria and Relevant Citations

| **Probiotic-Associated Bacteria** | **Citation(s)** |
| --- | --- |
| *Bifidobacterium adolescentis* | <https://pubmed.ncbi.nlm.nih.gov/25386066/> |
| *Bifidobacterium animalis* | <https://pubmed.ncbi.nlm.nih.gov/25386066/>  https://pubmed.ncbi.nlm.nih.gov/28114246/ |
| *Bifidobacterium bifidum* | [https://pubmed.ncbi.nlm.nih.gov/25386066/ https://pubmed.ncbi.nlm.nih.gov/22071814/](https://pubmed.ncbi.nlm.nih.gov/25386066/) |
| *Bifidobacterium breve* | <https://pubmed.ncbi.nlm.nih.gov/25386066/> |
| *Bifidobacterium longum* | [https://pubmed.ncbi.nlm.nih.gov/25386066/ https://pubmed.ncbi.nlm.nih.gov/22071814/](https://pubmed.ncbi.nlm.nih.gov/25386066/) |
| *Lactobacillus acidophilus* | <https://pubmed.ncbi.nlm.nih.gov/22071814/> |
| *Lactobacillus brevis* | <https://pubmed.ncbi.nlm.nih.gov/33169341/> |
| *Lactobacillus casei* | <https://pubmed.ncbi.nlm.nih.gov/22071814/> |
| *Lactobacillus delbrueckii* | <https://pubmed.ncbi.nlm.nih.gov/22071814/>  https://pubmed.ncbi.nlm.nih.gov/28114246/ |
| *Lactobacillus fermentum* | <https://pubmed.ncbi.nlm.nih.gov/31432401/> |
| *Lactobacillus gasseri* | <https://pubmed.ncbi.nlm.nih.gov/12182746/> |
| *Lactobacillus johnsonii* | <https://pubmed.ncbi.nlm.nih.gov/30580660/> |
| *Lactobacillus paracasei* | <https://pubmed.ncbi.nlm.nih.gov/32326347/> |
| *Lactobacillus plantarum* | <https://pubmed.ncbi.nlm.nih.gov/22071814/> |
| *Lactobacillus rhamnosus* | <https://pubmed.ncbi.nlm.nih.gov/22071814/> |
| *Lactobacillus reuteri* | <https://pubmed.ncbi.nlm.nih.gov/17032283/> |
| *Lactobacillus salivarius* | <https://pubmed.ncbi.nlm.nih.gov/24010610/> |
| *Stroptococcus thermophilus* | <https://pubmed.ncbi.nlm.nih.gov/22071814/>  https://pubmed.ncbi.nlm.nih.gov/28114246/ |
| *Bacteroides thetaiotaomicron* | <https://pubmed.ncbi.nlm.nih.gov/31324278/> |
| *Bacteroides fragilis* | <https://pubmed.ncbi.nlm.nih.gov/31324278/> |
| *Bacteroides ovatus* | <https://pubmed.ncbi.nlm.nih.gov/30459813/> |
| *Bacteroides vulgatus* | <https://pubmed.ncbi.nlm.nih.gov/34195297/> |
