## Supplementary figures and images for "Administration of *Bifidobacterium animalis* subsp. *lactis* Strain BB-12^®^ in Healthy Children: Characterization, Functional Composition, and Metabolism of the Gut Microbiome"

### Supplemental Figure 1

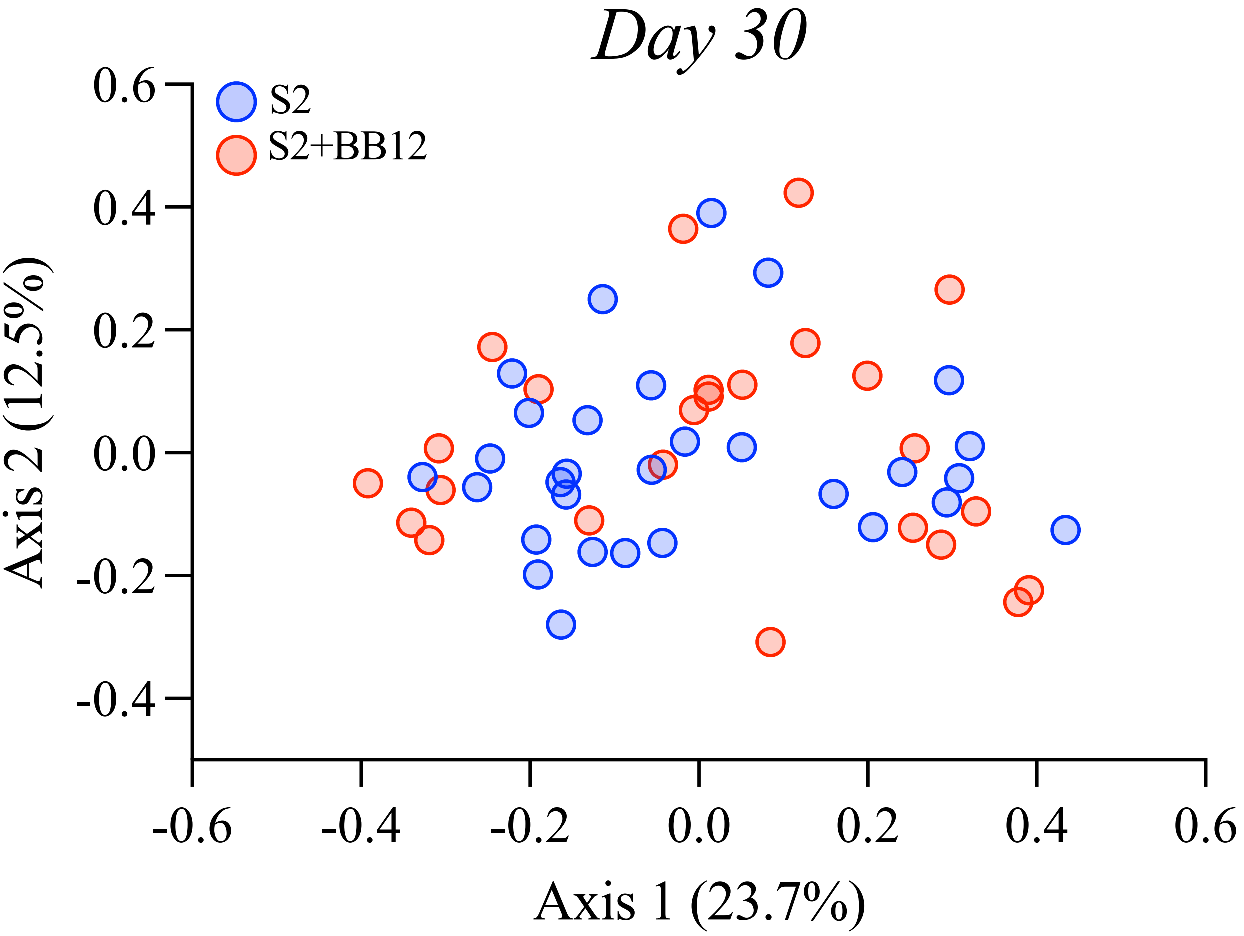

### Supplemental Figure 2

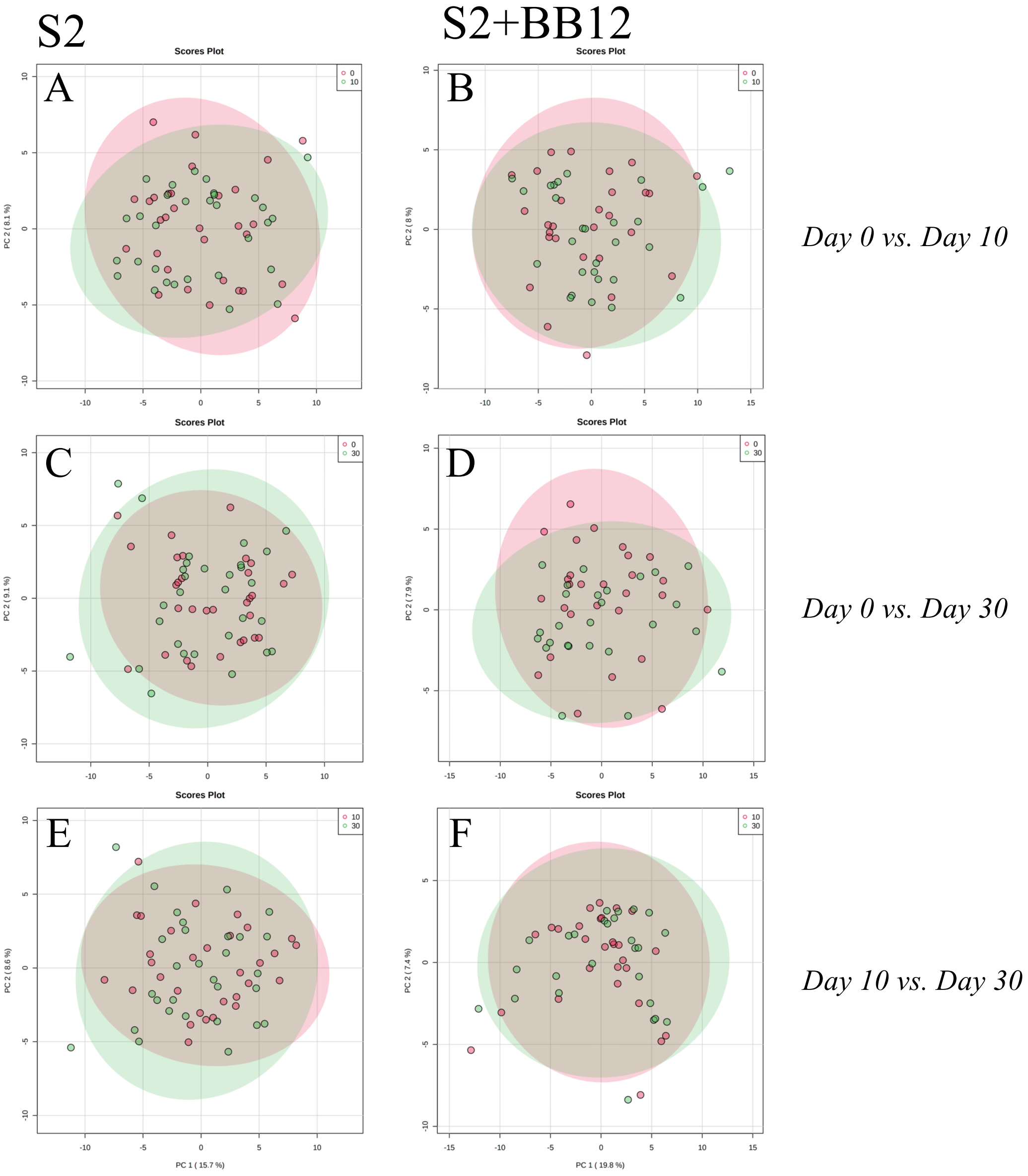

### Supplemental Table 3

**Supplemental Table 3**

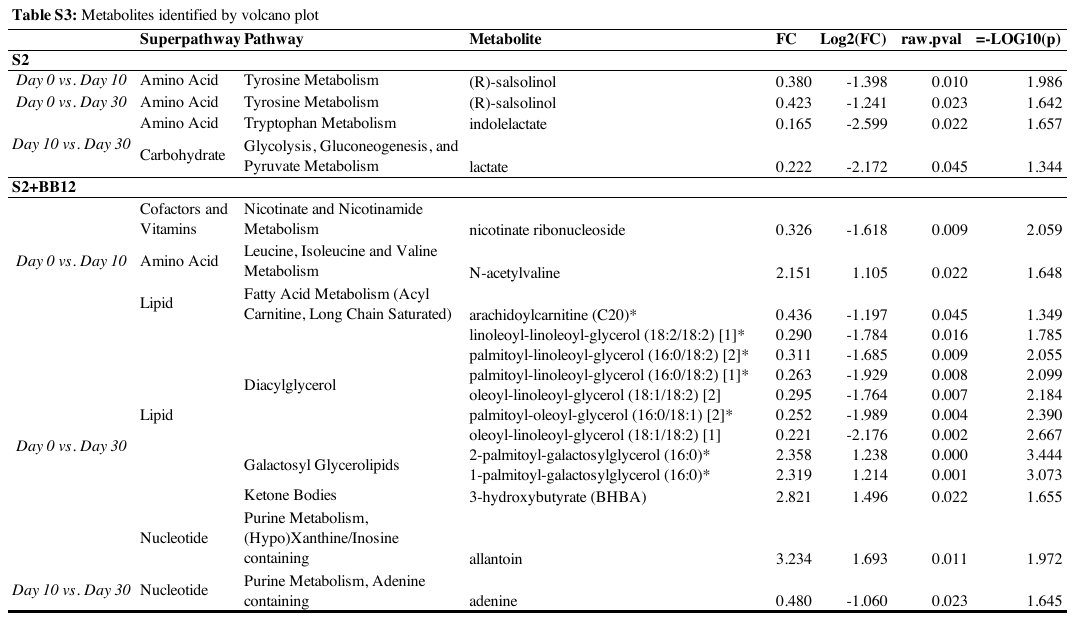
