## Supplemental Table 4 for "Administration of *Bifidobacterium animalis* subsp. *lactis* Strain BB-12^®^ in Healthy Children: Characterization, Functional Composition, and Metabolism of the Gut Microbiome"

**Supplemental Table 4. Selected Amino Acid-Associated Metabolites with Significant Model Effects**

| **Metabolite** | **Effect** | **DF** | **F Ratio** | ***P*** |
| --- | --- | --- | --- | --- |
| 3-methyl-2-oxobutyrate | Group | 1, 56 | 0.21 | 0.648 |
| 3-methyl-2-oxobutyrate | Time | 2, 109.74 | 5.78 | **0.004** |
| 3-methyl-2-oxobutyrate | Group * Time | 2, 109.74 | 1.74 | 0.179 |
| 3-methyl-2-oxovalerate | Group | 1, 58.40 | 2.98 | 0.089 |
| 3-methyl-2-oxovalerate | Time | 2, 111.81 | 3.82 | **0.025** |
| 3-methyl-2-oxovalerate | Group * Time | 2, 111.81 | 2.57 | 0.081 |
| 4-methyl-2-oxopentanoate | Group | 1, 58.77 | 3.43 | 0.068 |
| 4-methyl-2-oxopentanoate | Time | 2, 112.12 | 5.17 | **0.007** |
| 4-methyl-2-oxopentanoate | Group * Time | 2, 112.12 | 2.27 | 0.108 |
| Alanine | Group | 1, 57.34 | 3.00 | 0.088 |
| Alanine | Time | 2, 111.56 | 7.08 | **0.001** |
| Alanine | Group * Time | 2, 111.56 | 2.79 | 0.066 |
| Aspartate | Group | 1, 57.64 | 5.38 | **0.024** |
| Aspartate | Time | 2, 112.11 | 5.88 | **0.004** |
| Aspartate | Group * Time | 2, 112.11 | 0.008 | 0.992 |
| Cysteine s-sulfate | Group | 1, 57.52 | 3.96 | 0.051 |
| Cysteine s-sulfate | Time | 2, 111.21 | 4.97 | 0.009 |
| Cysteine s-sulfate | Group * Time | 2, 111.21 | 1.78 | 0.173 |
| Glutamate | Group | 1, 56.96 | 3.97 | 0.051 |
| Glutamate | Time | 2, 111.43 | 3.50 | 0.034 |
| Glutamate | Group * Time | 2, 111.43 | 0.98 | 0.380 |
| Glycine | Group | 1, 57.99 | 3.38 | 0.071 |
| Glycine | Time | 2, 111.93 | 8.76 | **<0.001** |
| Glycine | Group * Time | 2, 111.93 | 3.81 | **0.025** |
| Histidine | Group | 1, 57.21 | 1.74 | 0.193 |
| Histidine | Time | 2, 110.12 | 6.07 | **0.003** |
| Histidine | Group * Time | 2, 110.12 | 0.98 | 0.378 |
| Indolelactate | Group | 1, 59.98 | 1.17 | 0.284 |
| Indolelactate | Time | 2, 102.24 | 2.25 | 0.111 |
| Indolelactate | Group * Time | 2, 110.12 | 3.51 | **0.034** |
| Isoleucine | Group | 1, 58.18 | 3.26 | 0.076 |
| Isoleucine | Time | 2, 112.13 | 4.03 | **0.020** |
| Isoleucine | Group * Time | 2, 112.13 | 2.09 | 0.129 |
| Lysine | Group | 1, 58.35 | 10.07 | **0.002** |
| Lysine | Time | 2, 112.29 | 4.67 | **0.011** |
| Lysine | Group * Time | 2, 112.29 | 1.57 | 0.212 |
| Methionine | Group | 1, 58.21 | 4.79 | **0.033** |
| Methionine | Time | 2, 112.15 | 2.31 | 0.104 |
| Methionine | Group * Time | 2, 112.15 | 0.90 | 0.411 |
| N2-acetyllysine | Group | 1, 56.18 | 9.92 | **0.003** |
| N2-acetyllysine | Time | 2, 111.00 | 0.46 | 0.634 |
| N2-acetyllysine | Group * Time | 2, 111.00 | 0.71 | 0.494 |
| N-acetylasparagine | Group | 1, 57.72 | 7.43 | **0.008** |
| N-acetylasparagine | Time | 2, 110.24 | 4.25 | **0.017** |
| N-acetylasparagine | Group * Time | 2, 110.24 | 0.79 | 0.455 |
| N-acetylglutamine | Group | 1, 56.36 | 3.43 | 0.069 |
| N-acetylglutamine | Time | 2, 110.54 | 8.32 | **<0.001** |
| N-acetylglutamine | Group * Time | 2, 110.24 | 2.95 | 0.057 |
| N-acetylleucine | Group | 1, 57.89 | 0.97 | 0.329 |
| N-acetylleucine | Time | 2, 112.54 | 4.92 | **0.009** |
| N-acetylleucine | Group * Time | 2, 112.54 | 0.97 | 0.381 |
| N-acetylserine | Group | 1, 57.71 | 3.84 | 0.055 |
| N-acetylserine | Time | 2, 107.05 | 0.13 | 0.877 |
| N-acetylserine | Group * Time | 2, 107.05 | 3.11 | **0.049** |
| N-acetylthreonine | Group | 1, 57.76 | 0.66 | 0.421 |
| N-acetylthreonine | Time | 2, 111.28 | 4.82 | **0.010** |
| N-acetylthreonine | Group * Time | 2, 111.28 | 2.28 | 0.107 |
| N-butyryl-leucine | Group | 1, 57.57 | 6.30 | **0.015** |
| N-butyryl-leucine | Time | 2, 108.10 | 3.73 | **0.027** |
| N-butyryl-leucine | Group * Time | 2, 108.10 | 0.42 | 0.660 |
| Phenylalanine | Group | 1, 58.41 | 3.42 | 0.069 |
| Phenylalanine | Time | 2, 112.33 | 4.60 | **0.012** |
| Phenylalanine | Group * Time | 2, 112.33 | 2.30 | 0.105 |
| Pyroglutamine | Group | 1, 58.24 | 0.00 | 0.968 |
| Pyroglutamine | Time | 2, 112.35 | 2.30 | 0.105 |
| Pyroglutamine | Group * Time | 2, 112.35 | 5.18 | **0.007** |
| Serine | Group | 1, 57.70 | 5.30 | **0.025** |
| Serine | Time | 2, 111.71 | 5.41 | **0.006** |
| Serine | Group * Time | 2, 111.71 | 1.15 | 0.321 |
| Tryptophan | Group | 1, 58.23 | 3.95 | 0.051 |
| Tryptophan | Time | 2, 112.50 | 3.15 | **0.047** |
| Tryptophan | Group * Time | 2, 112.50 | 2.07 | 0.131 |
| Tyrosol | Group | 1, 58.89 | 4.60 | **0.036** |
| Tyrosol | Time | 2, 111.47 | 0.19 | 0.830 |
| Tyrosol | Group * Time | 2, 111.47 | 0.49 | 0.613 |
| Valine | Group | 1, 58.36 | 2.95 | 0.091 |
| Valine | Time | 2, 112.42 | 5.76 | **0.004** |
| Valine | Group * Time | 2, 112.42 | 2.10 | 0.127 |

Statistics performed on log-transformed data.
